# Distinct Somatic Mutational Landscapes in Paired Blood and Arterial Tissues in Coronary Artery Disease

**DOI:** 10.64898/2026.09.21.26363617

**Authors:** Zhi Chng Lau, Md Zanzibul Tareq, Kar Seng Sim, Xiao Yun Lin, Hao Wu, Arshia Naaz, Umamaheswari Muniasamy, Chiea Chuen Khor, Woon-Puay Koh, Vitaly A. Sorokin, Rajkumar Dorajoo

## Abstract

**Background:** Clonal hematopoiesis of indeterminate potential (CHIP) is associated with ageing and atherosclerotic coronary artery disease (CAD), but evidence has largely come from peripheral blood. Whether somatic mutational profiles differ between arterial tissue and blood, and whether hematopoietic clones are represented in arterial tissue, remain poorly understood.

**Methods:** We performed deep whole-exome sequencing on 302 blood granulocytes and 263 arterial tissues from patients undergoing coronary artery bypass graft surgery, including 221 matched blood-tissue pairs, with mean sequencing depths of 130.5X and 120.8X, respectively. Somatic variants were identified using Mutect2. Associations with age and CAD complexity, assessed using SYNTAX score, were evaluated using multivariable regression. Primary analyses focused on variants with variant allele fractions of 10-25%.

**Results:** CHIP was detected in 24/302 (7.9%) blood granulocytes, predominantly involving *DNMT3A*, *TET2*, and *ASXL1*. In contrast, somatic mutations in CHIP-associated genes in arterial tissues showed a distinct profile dominated by *CALR*, *SETDB1*, and *PDSS2*. Somatic mutation carrier status increased with age in blood (β = 0.07, *p* = 0.02) and arterial tissues (β = 0.06, *p* = 0.04). Blood CHIP was associated with lower SYNTAX score (β = −1.05, *p* = 2.66 x 10^-3^), partly driven by *DNMT3A* carriers (β = −1.06, *p* = 0.04), whereas arterial *CALR* mutations were associated with higher SYNTAX score (β = 1.14, *p* = 0.03). Among 221 matched pairs, only four identical variants, involving *CHEK2*, *DNMT3A*, *GNB1*, and *SRSF1*, were detected in both compartments. Genome-wide analyses showed compartment specificity, with bile acid metabolism enriched among arterial tissue-specific mutations (*P*_Adj_ = 0.038).

**Conclusion:** Blood and arterial tissues in CAD harbour distinct somatic mutational landscapes with limited cross-compartment sharing. These findings suggest that cardiovascular somatic mutations may reflect both hematopoietic and arterial tissue-associated processes and motivate cell-resolved and spatial studies to define their origins and roles in cardiovascular ageing.

## Introduction

Ageing is accompanied by the lifelong accumulation of somatic mutations that progressively reshape the genomic landscape of individual tissues. While most somatic mutations are functionally neutral, a subset confers selective advantages that promote clonal expansion, contributing to tissue dysfunction and age-related diseases. Increasing evidence suggests that somatic evolution represents a fundamental biological process of ageing, linking genomic instability with chronic inflammation, impaired tissue homeostasis and declining organ function [1, 2]. However, the extent to which somatic mutational processes differ across human tissues and contribute to organ-specific ageing remains poorly understood.

Clonal hematopoiesis of indeterminate potential (CHIP) has emerged as one of the best-characterized examples of age-associated somatic evolution in humans. CHIP is defined by the expansion of hematopoietic stem cell clones carrying acquired driver mutations, most commonly in *DNMT3A*, *TET2*, and *ASXL1*, and increase markedly with advancing age. Beyond its established association with hematological malignancies, CHIP has been consistently linked to cardiovascular disease (CVD), heart failure, stroke, and all-cause mortality [3, 4, 5, 6]. Experimental studies have demonstrated that CHIP-associated mutations enhance inflammatory activation of myeloid cells, promoting inflammasome signaling and accelerating atherosclerosis, establishing CHIP as a major mechanistic link between ageing and CVD [1, 2, 7].

Despite these advances, current understanding of CHIP is derived almost exclusively from analyses of peripheral blood. Consequently, it remains unclear whether the cardiovascular consequences of CHIP primarily reflect systemic inflammatory effects of circulating mutant immune cells or whether these clones are directly represented within ageing vascular tissues. Furthermore, little is known about whether blood and vascular tissues undergo similar or distinct patterns of somatic evolution during ageing. Recent studies have identified somatic mutations within atherosclerotic lesions and vascular tissues, suggesting that clonal expansion is not restricted to the hematopoietic compartment [8, 9]. However, direct comparisons of somatic mutational landscapes between matched blood and vascular tissues in humans remain scarce, limiting our understanding of tissue-specific somatic evolution.

Coronary artery disease (CAD) provides a unique human model to investigate these questions because both peripheral blood and disease-relevant vascular tissues can be obtained from patients undergoing coronary artery bypass graft (CABG) surgery. Simultaneous characterization of somatic mutations across these compartments enables evaluation of whether age-associated hematopoietic clones are represented within vascular tissue and whether distinct somatic mutational processes contribute to vascular pathology.

Here, we performed deep whole-exome sequencing of peripheral blood granulocytes (n=302) and arterial tissues (n=263), with mean sequencing depth of 130.5X and 120.8X respectively, including approximately 221 matched blood-tissue pairs, from Chinese male patients who underwent CABG surgery. We first characterized age-associated CHIP in blood and then systematically compared somatic mutational landscapes between blood and arterial tissues. Finally, we investigated the extent to which blood CHIP mutations were detectable within matched vascular tissue and explored the relationships between compartment-specific somatic mutations and coronary disease severity.

## Methods

### Study subjects

We recruited a total of 302 CAD patients who underwent CABG surgery at the National University Hospital of Singapore between 2010 and 2014 [10]. All patients provided consent to participate by donating blood immediately prior to surgery and by donating arterial tissue samples harvested during the procedure. This study was approved by the National Healthcare Group (NHG) Domain Specific Review Board (NHG DSRB Reference Number: 2009/00216). Written informed consent was obtained from each patient before enrollment (NHG Tissue Bank Registration Number: NUH/2009-0073). The study protocol adhered to the ethical guidelines of the 1975 Declaration of Helsinki.

### Sample processing

Whole blood was collected pre-operatively from each patient, washed in phosphate-buffered saline and cold ammonium chloride solution (Stemcell Technologies), and processed by density gradient centrifugation. The supernatant was discarded, and the granulocyte pellet was resuspended in cell freezing medium. Arterial tissues were collected from the same patients during CABG surgery at the site of proximal anastomosis between the aorta and saphenous vein grafts [10]. Tissue punches were taken from non-atheromatous arterial walls in the proximal region of the ascending aorta, which shares an embryological origin with the coronary arteries. Samples were cryopreserved on dry ice immediately and subsequently stored in a liquid nitrogen tank.

Genomic DNA was extracted from arterial tissues and granulocyte samples using the Monarch^®^ Genomic DNA Purification Kit (New England Biolabs) according to the manufacturer’s instructions. DNA concentrations were determined using a DS-11 Spectrophotometer (DeNovis Inc.).

### Whole-exome sequencing and germline quality control

Whole-exome sequencing (WES) libraries were prepared using the Twist Exome 2.0 plus Comprehensive Exome Spike-in enrichment kits and sequenced on the DNBSEQ-T7 platform to an average sequencing depth of 130.5X in blood granulocytes (n=302) and 120.8X in arterial tissues (n=263).

Raw paired-end reads were aligned to the human reference genome (hg19) using the Burrows-Wheeler Aligner (BWA) software packages [11], followed by duplicate removal using Picard (https://broadinstitute.github.io/picard/). Local realignment around known insertions and deletions and base quality score recalibration (BQSR) was subsequently performed using the Genome Analysis Toolkit (GATK) software [12]. Variant calling on the mapped reads was performed for each sample using GATK HaplotypeCaller to generate genomic variant call format (gVCF) files. The gVCF files from all samples were then jointly called, followed by filtering on the called genotypes using variant quality score recalibration (VQSR) at a threshold of 99.5% for both SNP and INDEL variants using GATK.

Genotype-level and variant-level quality control (QC) were performed. Individual genotype calls with genotype quality (GQ) < 20 or read depth (DP) < 10 were set as missing, while variants with a mean DP <10 across all samples were excluded. Additional sample- and variant-level QC included sample call rate > 90%, variant call rate > 90%, extreme deviation from Hardy-Weinberg equilibrium *P_HWE_* > 1×10^-10^, identity-by-descent (IBD) analysis, and principal component analysis (PCA). No samples were excluded on the basis of sample call rate. PCA identified and excluded six outlier samples, comprising two blood granulocytes and four arterial wall tissue samples. IBD analysis confirmed 221 blood-tissue pairs as genetically matched samples, which were retained for downstream paired analyses.

### Somatic variant calls, variant annotation and filtering

For somatic mutation detection, the BAM files generated on each sample following BQSR were independently analyzed using Mutect2 implemented in GATK [13, 14]. Mutect2 identifies genomic sites with evidence of sequence variation and performs local haplotype reassembly to improve variant detection. To reduce recurrent sequencing artifacts, Mutect2 was run with a panel of normals constructed from WES data of 395 unrelated East Asian individuals aged 10-18 years. Standard Mutect2 processing and filtering procedures were applied to identify and remove potential sequencing and technical artifacts, and only variants designated “PASS” in the resulting VCF files were retained for downstream analyses. Variants identified in the variant allele fraction (VAF) range between 2% to 25% were extracted. The lower threshold enabled sensitivity analyses including smaller clonal populations, consistent with previous CHIP studies [14], and the upper threshold of 25% was utilized to limit heterozygous germline calls. Primary analyses focused on variants with VAFs between 10% and 25%, representing larger somatic clones.

Identified somatic variants were functionally annotated using the Ensembl Variant Effect Predictor (VEP) version 110.1. Downstream analyses were restricted to variants predicted to have a high or moderate functional impact on the protein product. This retained canonical splice site mutations (splice acceptor and donor variants) alongside other highly disruptive alterations like frameshift, stop-gained, and missense mutations. Synonymous, intronic, untranslated region, and intergenic variants were excluded. To limit germline variants, variants with an observed minor allele frequency > 0.1% in any reference population from the large-scale GnomAD database were excluded [15].

All data analytics for sensitive human sequencing data were performed in the Research Assets Provisioning and Tracking Online Repository (RAPTOR) [16].

### CHIP carriers

We identified CHIP carriers on the basis of prespecified list of variants in 95 genes known to be recurrent drivers of myeloid malignancies (Supplementary Table 2) [14, 17]. Primary analysis was based on variants with VAF threshold of 10-25% to enrich large somatic clones that are more consistently detected across population-based CHIP studies while minimizing potential inclusion of heterogenous germline variants. A broader VAF threshold of 2-25% was evaluated in sensitivity analyses to assess whether inclusion of smaller clonal populations altered the observed associations.

### SYNTAX scores

Coronary angiography provides detailed anatomical assessment of CAD, and the SYNTAX score (SS), developed by the *SYN*ergy between percutaneous coronary intervention with *TAX*us and cardiac surgery (SYNTAX) trial, is a validated measure of coronary lesion complexity and disease severity. The SS was calculated blindly by independent medical staff member using the official online calculator (https://syntaxscore.org/). The scores were categorized into three categories: low SS (<23), intermediate SS (23 – 32), high SS (> 32). The SS were rank inverse normalized before performing statistical analyses.

### Statistical Analyses

All the statistical analyses were performed with the use of the R statistical package (www.r-project.org). Continuous phenotypes such as age and SS were rank-based inverse normal transformed respectively before regression analyses. Participants were classified as CHIP carriers if at least one somatic mutation in the curated CHIP gene panel was detected at a VAF threshold of 10-25%, and as non-CHIP carriers otherwise. Associations between CHIP carrier status with age and SS were evaluated using multivariable linear regression, with diabetes, hypertension, hyperlipidemia, and smoking status included as covariates. Additionally, we evaluated the association with SS categories using proportional odds ordinal logistic regression, with adjustment for age, smoking, diabetes, hypertension, hyperlipidemia, and smoking status.

To determine whether individual CHIP genes contributed to the observed phenotype associations, gene-level analyses were performed separately for each mutated CHIP gene. Patients carrying a mutation in the gene were compared with patients without detectable CHIP mutations. For each gene, continuous outcomes (age, SS) were analyzed using similar multivariable linear regression. Two-sided *P* values less than 0.05 were considered statistically significant for prespecified CHIP analyses. Gene-level analyses for individual somatic mutations to clinical phenotypes were considered exploratory and interpreted using nominal *P* values.

### Gene-set enrichment analysis

Gene-set enrichment analysis was performed using FUMA (Functional Mapping and Annotation) to investigate the biological pathways represented by genes that carried somatic mutations. We queried the default curated gene sets from the Molecular Signatures Database (MSigDB). To avoid the redundant pathways and focus on robust biological signals, the gene enrichments were based on the 50 distinct Hallmark gene sets. For the primary analysis, genes with somatic mutations uniquely detected in at least 10 carriers in arterial tissues were included. As a sensitivity analysis, a less stringent threshold of at least three carriers was applied to assess the robustness of the pathway enrichment findings. Multiple testing correction was applied using the Benjamini-Hochberg false discovery rate (FDR), with significance defined at an adjusted *P* value < 0.05.

## Results

### Classical CHIP landscape was observed in peripheral blood of CAD patients

The clinical characteristics of the 302 CAD patients who underwent CABG surgery at the National University Hospital of Singapore between 2010 and 2014 were presented in Supplementary Table 1. Briefly, these patients comprised Chinese men with a median age of 62 years (interquartile range [IQR], 57 – 67) and a high prevalence of cardiovascular risk factors, including type 2 diabetes (53.3%), smoking (57.4%), hypertension (81.1%), and hyperlipidemia (97.7%). Using a VAF threshold of 10-25%, 24 patients carried canonical CHIP mutations in peripheral blood (Figure 1). The most frequently mutated genes were *DNMT3A*, followed by *TET2*, and *ASXL1*, consistent with previous data reported as the most commonly mutated genes linked to clonal hematopoiesis. CHIP prevalence increased significantly with advancing age (β = 0.068, *p* = 0.018) (Table 1), supporting the validity of the variant-calling strategy and demonstrating that the mutational landscape in this Chinese CAD cohort closely resembles that reported in predominantly European populations.

**Figure 1.**
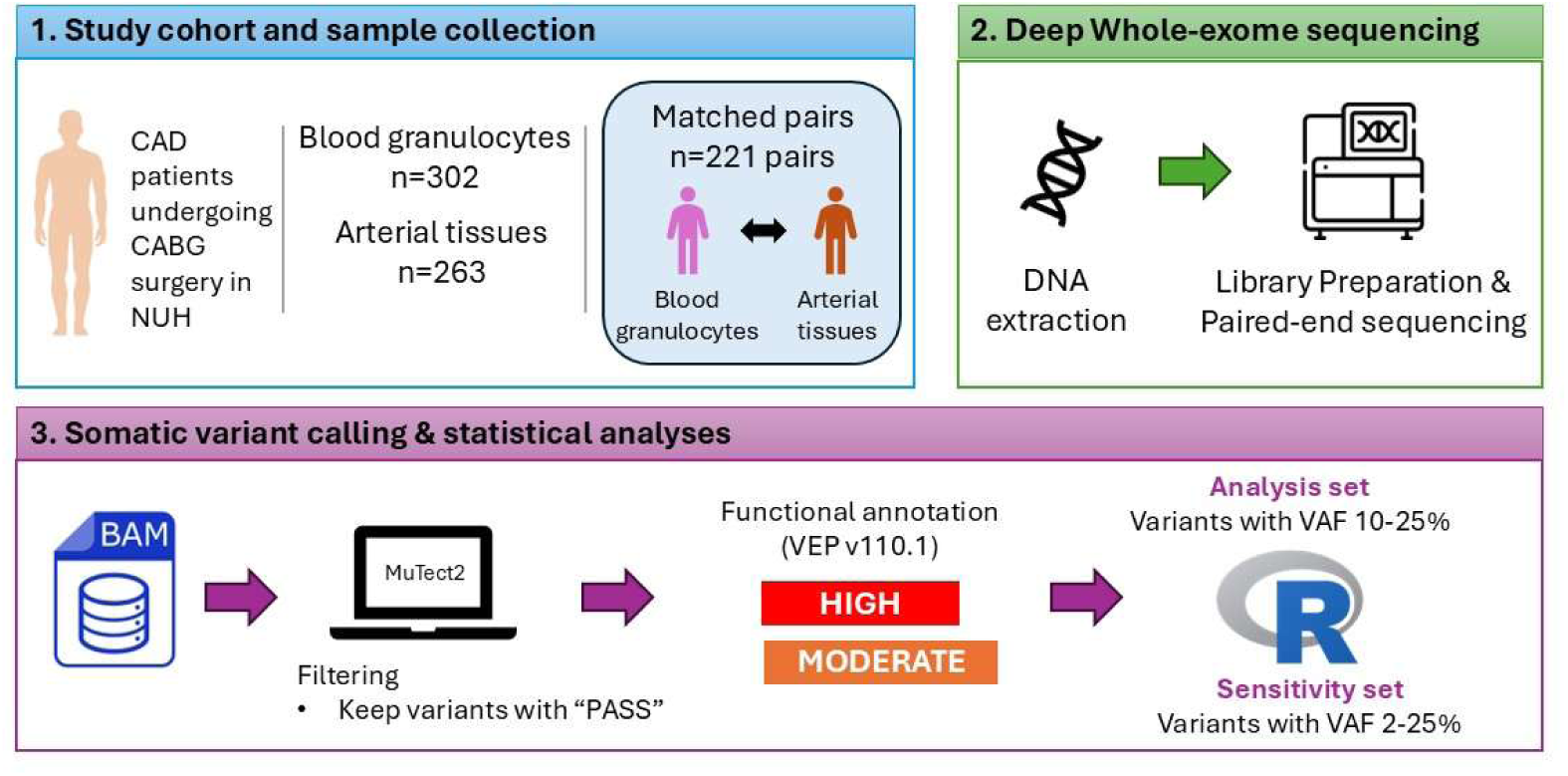
Study design and analytical workflow for somatic mutation profiling in blood granulocytes and arterial tissues. Patients with coronary artery disease (CAD) undergoing coronary artery bypass grafting (CABG) surgery at the National University Hospital (NUH) were included in the study. Blood granulocytes (n=302) and arterial tissues (n=263) underwent deep whole-exome sequencing, including 221 matched blood-arterial tissue pairs. Following DNA extraction, library preparation and paired-end sequencing, somatic variants were called independently from BAM files using Mutect2. Only variants passing Mutect2 filtering were retained and functionally annotated using Ensembl Variant Effect Predictor (VEP) v110.1. Downstream analyses were restricted to variants predicted to have HIGH or MODERATE functional impact. Primary analysis included variants with variant allele fraction (VAF) threshold of 10-25%, while a broader VAF range of 2-25% was evaluated in sensitivity analyses. Statistical analyses were performed in R. BAM, Binary alignment map; CABG, Coronary artery bypass graft; CAD, coronary artery disease; NUH, National University Hospital; VAF, variant allele fraction; VEP, variant effect predictor.

**Figure 2.**
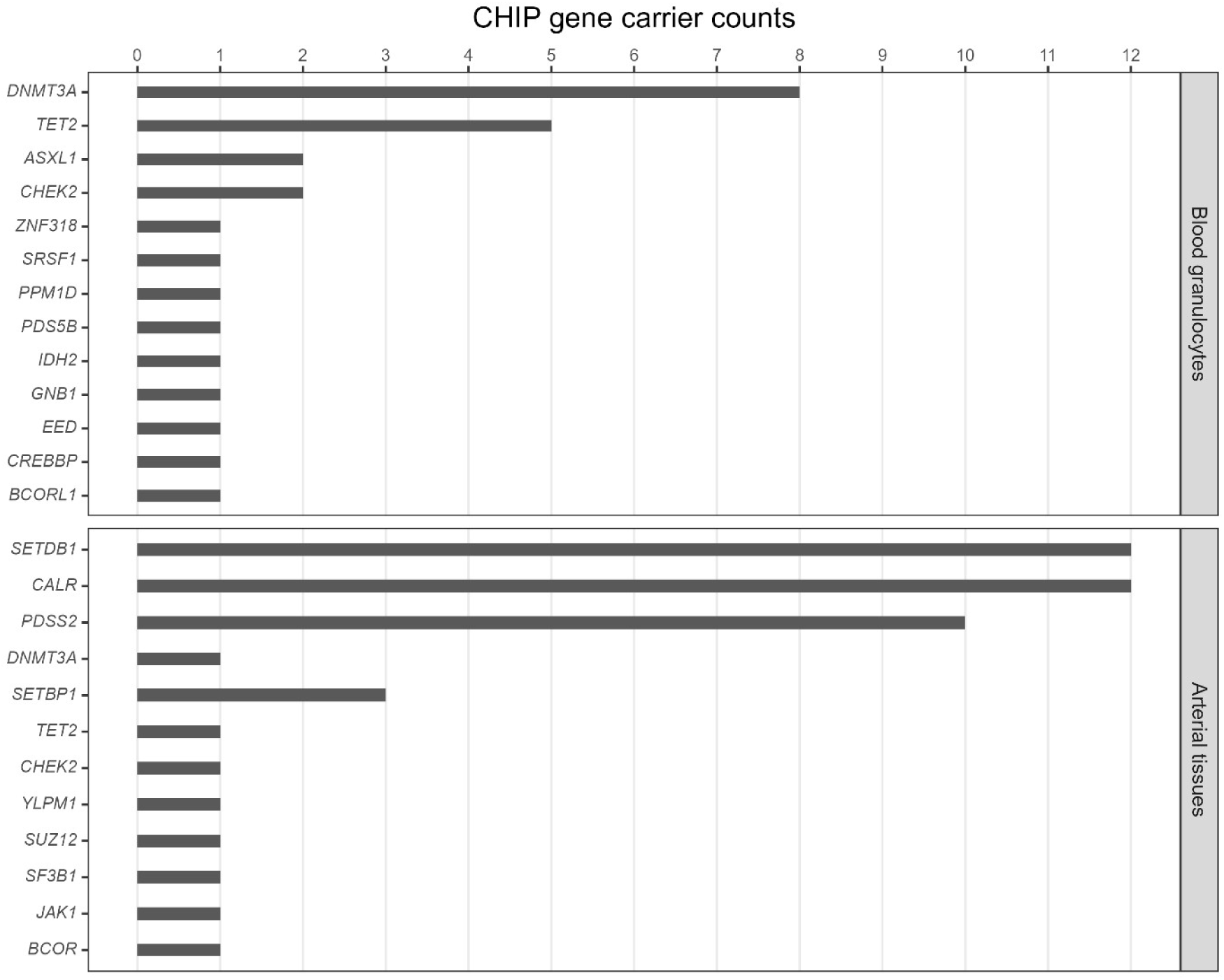
Distribution of somatic mutations in CHIP-associated genes in blood granulocytes and arterial tissues at variant allele fraction (VAF) threshold of 10-25%. Carrier counts for somatic mutation in CHIP-associated genes detected in blood granulocytes (upper panel) and arterial tissues (lower panel) among patients with coronary artery disease. Bars indicate the number of carriers harbouring a somatic mutation in each gene. Genes with at least one carrier are shown.

**Table 1.** Shared CHIP variants between matched blood granulocytes and arterial tissues. Identical CHIP variants detected in both blood granulocytes matched arterial tissues are shown with chromosome position, reference and alternate alleles, allele depth, and variant allele fraction (VAF) in each tissue. Chr, chromosome; REF, reference allele, ALT; alternate allele; VAF, variant allele fraction

| Gene | Chr | Position | REF | ALT | Granulocytes | Granulocytes | Arterial Tissues | Arterial Tissues |
| --- | --- | --- | --- | --- | --- | --- | --- | --- |
|  |  |  |  |  | Ref, Alt reads | VAF | Ref, Alt reads | VAF |
| *CHEK2* | 22 | 29099513 | ATCT | A | 52,16 | 0.235 | 61,10 | 0.141 |
| *DNMT3A* | 2 | 25463575 | G | C | 72,6 | 0.077 | 132,12 | 0.083 |
| *GNB1* | 1 | 1747229 | T | C | 134,17 | 0.113 | 101,7 | 0.065 |
| *SRSF1* | 17 | 56084319 | C | G | 129,42 | 0.246 | 140,15 | 0.097 |

### Arterial tissue exhibited a distinct somatic mutation landscape

In contrast to peripheral blood, arterial tissue demonstrated a markedly different distribution of somatic mutations. Somatic mutations in CHIP-associated genes were detected in arterial tissue from 25 patients (Figure 1). The most commonly mutated genes were *CALR*, *SETDB1* and *PDSS2*. Similar to peripheral blood, carriers of somatic mutations in CHIP-associated genes in the arterial tissues were also associated with increased age (β = 0.063, *p* = 0.041) (Table 1).

To characterize the broader landscape of somatic mutations beyond established CHIP genes, we expanded the analysis to include all genes harbouring somatic variants with VAF threshold of 10-25%. Distinct tissue-specific patterns were observed between the arterial tissues and blood granulocytes (Supplementary Table 3). Among 1,935 genes with mutations detected within this VAF range, 46.2% (894 genes) were detected in one or more arterial tissues but were not detected in blood granulocytes. Conversely, 37.6% (727 genes) were detected exclusively in blood granulocytes but not in the arterial tissues. However, these blood-specific mutations generally occur at low frequencies, with a maximum of five carriers. The *MVP* gene was the most frequently mutated blood-specific gene, occurring in five carriers while not detected in the arterial tissue.

To investigate the biological pathways represented by somatic mutations in the arterial tissues, we performed gene-set enrichment analysis. Genes with somatic mutations detected in at least ten carriers in arterial tissues (VAF threshold of 10-25%) showed significant enrichment for bile acid metabolism pathway (nominal *P* = 7.68 x 10^-4^; *P_Adj_* = 0.038), contributed by *ALDH8A1*, *ABCA3*, and *MLYCD* (Supplementary Table 4). In a sensitivity analysis, we included somatic genes detected in at least three carriers in arteries. Bile acid metabolism remained significantly enriched (nominal *P* = 1.09 x 10^-4^; *P_Adj_* = 5.43 x 10^-3^), with six overlapping genes (*SULT1B1*, *BMP6*, *ALDH8A1*, *APOA1*, *ABCA3*, and *MLYCD*), supporting the robustness of this finding. Collectively, the enrichment of metabolism gene set across VAF thresholds suggests that somatic mutations in arterial tissues may involve genes participating in lipid and bile-acid metabolic pathways.

### Blood and arterial tissues mutations showed limited overlap

Among 221 genetically matched blood-arterial tissue pairs, 27 patients carried canonical CHIP mutations in blood (VAF 2-25%). Nineteen distinct CHIP variants were identified in these patients, of which four identical variants (21.1%) were detected in both matched blood and arterial tissue from the same individuals. This observed overlap was greater than expected by chance (Binomial *P* = 0.011). Shared variants involved *DNMT3A*, *CHEK2*, *GNB1*, and *SRSF1* (Table 1). For three variants (at *CHEK2*, *GNB1*, and *SRSF1*), VAFs were generally lower in arterial tissue than in blood, suggesting partial representation of hematopoietic clones within the vascular compartment. A single *DNMT3A* mutation exhibited comparable VAF in both compartments.

### Exploratory evaluations with coronary disease complexity

We next explored whether compartment-specific somatic mutations were associated with anatomical severity of CAD. SS were available for 157 of 302 patients (52.0%), including 26 patients (16.6%) with low SS, 71 patients (45.2%) with intermediate SS, and 60 patients (38.2%) with high SS (Supplementary Table 1). Peripheral blood CHIP was associated with lower SS (β = −1.046, *p* = 2.66 x 10^-3^), an observation driven by *DNMT3A* mutations (β = −1.062, *p* = 0.039) (Table 2). Consistently, ordinal logistic regression of SS categories (low, intermediate, and high) showed that blood CHIP carriers had substantially lower odds of being in a higher SYNTAX category (OR = 0.08, 95% CI 0.02 – 0.33; *p* = 4.36 x 10^-4^). Conversely, somatic mutations in CHIP-associated genes identified in arterial tissues were not associated with either SS (β = 0.257, *p* = 0.531) or SS category (OR = 1.18, 95% CI 0.26 – 5.30; *p* = 0.832). Nonetheless, somatic mutations in CHIP-associated genes identified in arterial tissue, specifically the *CALR* mutation, was associated with higher SS (β = 1.138, *p* = 0.033).

**Table 2.** Associations of overall CHIP and individual CHIP genes with age and SYNTAX scores. Multivariable regression analyses were performed among patients carrying somatic mutations with a VAF threshold of 10-25%. For overall CHIP analyses, patients carrying at least one CHIP mutation were compared with patients without detectable CHIP mutations. For gene-specific analyses, carriers of the indicated CHIP gene were compared with patients without detectable CHIP mutations; carriers of mutations in other CHIP genes were excluded. Age and SYNTAX scores were rank-based inverse normal transformed respectively before multivariable linear regression. SYNTAX scores were categorized as low, intermediate, and high and analyzed using ordinal logistic regression, with increasing category corresponding to greater CAD severity. All models were adjusted for age, smoking status, diabetes mellitus, hypertension, and hyperlipidemia, as appropriate. Values are presented as regression coefficients (β), standard errors (SE), odds ratio (OR), 95% confidence interval (95% CI), and two-sided *P*-values. CHIP, clonal hematopoiesis of indeterminate potential; VAF, variant allele fraction; SYNTAX, Synergy Between Percutaneous Coronary Intervention with TAXUS and Cardiac Surgery.

| Tissue |  | n | Outcome | β | SE | P value |
| --- | --- | --- | --- | --- | --- | --- |
|  |  | (Carrier vs non-CHIP) |  |  |  |  |
| Blood granulocytes | Overall | 24 vs 274 | Age | 0.068 | 0.029 | 0.018 |
|  |  | 9 vs 148 | SYNTAX score | -1.046 | 0.342 | 2.66 x 10<sup>-3</sup> |
|  |  |  |  | **OR** | **95% CI** |  |
|  |  | 9 vs 148 | SYNTAX category | 0.084 | 0.02 – 0.33 | 4.36 x 10<sup>-4</sup> |
|  |  |  |  | **β** | **SE** |  |
|  | *DNMT3A* | 8 vs 278 | SYNTAX score | -1.062 | 0.511 | 0.039 |
| Arterial tissues | Overall | 24 vs 236 | Age | 0.063 | 0.031 | 0.041 |
|  |  | 7 vs 125 | SYNTAX score | 0.257 | 0.409 | 0.531 |
|  |  |  |  | **OR** | **95% CI** |  |
|  |  | 7 vs 125 | SYNTAX category | 1.177 | 0.26 – 5.30 | 0.832 |
|  |  |  |  | **β** | **SE** |  |
|  | *CALR* | 12 vs 238 | SYNTAX score | 1.138 | 0.527 | 0.033 |

Sensitivity analyses using a broader VAF threshold of 2-25% yielded generally consistent findings (Supplementary Figure 1, Supplementary Table 5), including the compartment-specific mutation spectrum and age associations in both tissues. The association between arterial *CALR* mutations and SS was also retained with the broader VAF threshold, with *CALR* carriers exhibiting higher SS (β = 1.158, *p* = 0.030).

## Discussion

Deep whole-exome sequencing of blood granulocytes and arterial tissues from patients undergoing CABG surgery showed that age-associated somatic mutational landscapes differed substantially between hematopoietic and arterial compartments. While canonical CHIP spectrum dominated by *DNMT3A*, *TET2*, and *ASXL1* were observed in blood, a distinct distribution of recurrent somatic mutations was observed in arterial tissue. The limited sharing of identical variants across paired samples of blood and arterial tissues suggested that most CHIP variants detected in blood were not widely represented in arterial tissues, suggesting that accumulation of somatic mutations in CAD could differ by compartments.

Our findings in blood granulocytes concurred with other studies showing that CHIP prevalence increased with age, and the predominance of *DNMT3A*, *TET2*, and *ASXL1* was consistent with the established spectrum of age-related clonal hematopoiesis [3, 4]. These genes regulate epigenetic and hematopoietic processes, and mutations affecting them can promote clonal expansion and alter inflammatory functions of myeloid cells. Hence, CHIP has consequently emerged as an important link between ageing, inflammation, and CVD [1]. The consistency of these established features in our blood granulocytes samples supports the biological validity of the somatic mutation profiles identified in this CAD cohort and provides a reference against which the markedly different arterial mutational landscape may be interpreted.

In contrast, we showed a distinct spectrum of somatic mutations in CHIP-associated genes in arterial tissues, with *CALR*, *SETDB1*, and *PDSS2* among the most frequently affected genes. These genes were uncommon or absent among blood CHIP carriers in the same cohort, suggesting that the arterial somatic mutation landscape could not be explained solely by circulating hematopoietic clones.

The extension of analysis to all genome-wide somatic mutations reinforced this compartment-specific pattern and showed *MVP* (major vault protein) as the most frequently mutated gene in the blood. Experimental evidence implicates *MVP* in macrophage inflammatory signaling and plaque stability, with myeloid *MVP* promoting MMP-9 expression and plaque destabilization [18]. Thus, the identification of somatic *MVP* mutations is biologically intriguing, but their low frequency warrants cautious interpretation and additional experimental validation.

Pathway enrichment analysis of somatically mutated genes in arterial tissue identified bile acid metabolism as a significantly enriched pathway, with *ALDH8A1*, *ABCA3*, and *MLYCD* contributing to the enrichment. Previous studies have consistently reported disturbances in bile acid metabolism in CAD [19, 20]. Our findings provided a tissue-level genomic observation that raised the possibility that metabolic pathways might be represented in the arterial somatic mutational landscape. However, whether these somatic alterations directly influence related lipid metabolism remain unclear and warrants additional validation in independent cohorts.

The associations with CAD severity in our study provided further evidence that the biological significance of somatic mutations may depend on both gene identity and tissue compartment. Although *DNMT3A*-CHIP has been associated with adverse cardiovascular outcomes [21], the lower SS observed among *DNMT3A* carriers compared to non-carriers in our study may indicate that anatomical disease complexity and CHIP-associated inflammatory processes may capture different dimensions of cardiovascular risk rather than opposing biological effects. The SS primarily reflects the anatomical extent and complexity of coronary lesions, whereas previous studies have demonstrated that *DNMT3A*-mutant hematopoietic cells exhibit enhanced inflammatory signaling, including increased expression of IL-1β and IL-6 and activation of immune-cell responses [22]. Additionally, aortic vascular smooth muscle cells from CAD patients with lower SS exhibited a pro-inflammatory phenotype, including increased IL-1β expression at both the transcript and protein levels [23]. Thus, the lower SS observed among *DNMT3A* carriers may potentially reflect a greater contribution of inflammatory processes to cardiovascular risk that may not be necessarily accompanied by greater anatomical coronary disease complexity.

Conversely, *CALR* mutations detected in arterial tissue were associated with higher SS. *CALR* is strongly linked to myeloproliferative biology, and pathogenic *CALR* mutations can activate JAK-STAT signaling and promote abnormal hematopoietic proliferation [24, 25]. An association between *CALR* and CAD severity is therefore biologically plausible, but the cellular origin of *CALR* mutations detected in bulk arterial tissue remains unclear. The signal could arise from infiltrating hematopoietic cells, locally expanded vascular or stromal cells, or a mixture of cell populations. Consequently, the contrasting *DNMT3A* and *CALR* associations are best viewed as evidence that gene identity and tissue compartment may matter, rather than as proof of distinct causal CAD subtypes.

The paired design of our study enabled direct assessment of whether somatic variants detected in blood were also present in arterial tissue from the same individuals. Only four identical CHIP variants were shared across matched compartments; however, this overlap was greater than expected by chance, suggesting limited but non-random sharing of somatic variants between blood and arterial tissue. For most of the shared variants, VAFs were lower in arterial tissues than in blood, consistent with the presence of mutant hematopoietic cells within a heterogeneous vascular-tissue sample. This interpretation is supported by studies of peripheral arterial disease reporting CHIP mutations in both blood and atherosclerotic femoral lesions [9], as well as evidence of somatically expanded cell populations within end-stage atherosclerotic plaques [8]. Importantly, however, shared variants represented only a small fraction of the somatic mutations detected in either compartment, with blood and arterial tissue otherwise exhibiting largely distinct mutational profiles within the same individuals. Together, these findings suggest that somatic mosaicism in cardiovascular ageing may comprise both hematopoietic clones represented within vascular tissue and additional compartment-associated mutational processes. Bulk-tissue sequencing cannot distinguish whether arterial mutations arise from infiltrating hematopoietic cells, vascular-resident cells, or other cell populations. Cell-resolved and spatial sequencing will therefore be important to establish the cellular origins of these mutations and determine whether the distinct mutational profiles observed in arterial tissue reflect local clonal expansion or other vascular ageing processes.

This study has several strengths. Blood and arterial tissues were obtained from the same CAD patients, enabling simultaneous evaluation of somatic mutations across compartments. The high sequencing depth enabled sensitive detection of lower-VAF variants, which we explored in sensitivity analyses using a 2-25% VAF threshold.

Nevertheless, several limitations should also be considered. The cohort was modest in size, and analyses were based on small numbers of variant carriers. Bulk whole-exome sequencing may not provide cell-type and spatial resolution, thus limiting the determination of whether arterial tissues mutations originated from vascular-resident cells or infiltrating hematopoietic cells. The cross-sectional design precludes establishing temporal or causal relationships between somatic mutations and CAD progression. All participants were Chinese men with advanced CAD requiring CABG surgery, limiting generalizability to women, other ethnicities, and earlier stages of CAD. Lastly, arterial tissue was obtained from the proximal ascending aorta rather than coronary atherosclerotic plaque, and therefore represented vascular tissue adjacent to, rather than within, coronary lesions. Nevertheless, the study enabled an opportunity to investigate somatic mutations within the broader vascular compartment, allowing a clearer assessment of tissue-associated somatic mutation accumulation and the extent to which blood-derived CHIP variants are represented outside focal plaques.

In conclusion, our findings broaden the study of somatic mosaicism in cardiovascular disease beyond circulating hematopoietic cells and raise the possibility of arterial tissue-associated somatic processes in CAD. The enrichment of *CALR* mutations and bile acid metabolism pathways in arterial tissue provides hypotheses for vascular-specific mechanisms that warrant validation in independent cohorts. Future cell-resolved, spatial, and functional genomic studies are needed to establish the cellular origins of these mutations and determine their biological significance and potential contribution to cardiovascular ageing and atherosclerosis.

## Competing interests

The authors declare no competing interests.

## Authors’ contributions

Zhi Chng Lau: Conceptualization, Methodology, Validation, Formal analysis, Investigation, Visualization, Project administration, Writing – original draft, Writing – review & editing. Md Zanzibul Tareq: Formal analysis, Investigation, Writing – original draft, Writing – review & editing. Kar Seng Sim: Methodology, Formal analysis, Investigation, Writing – original draft, Writing – review & editing. Xiao Yun Lin: Investigation, Writing - review & editing. Hao Wu: Formal analysis, Visualization, Writing – review & editing. Arshia Naaz: Investigation, Writing - review & editing. Umamaheswari Muniasamy: Methodology, Writing – review & editing. Chiea Chuen Khor: Methodology, Writing – review & editing. Woon-Puay Koh: Supervision, Writing – original draft, Writing – review & editing. Vitaly A. Sorokin: Conceptualization, Supervision, Writing – original draft, Writing – review & editing. Rajkumar Dorajoo: Conceptualization, Formal analysis, Methodology, Validation, Supervision, Funding acquisition, Writing – original draft, Writing – review & editing.

## Funding sources

This study was supported by National University of Singapore Start-up Grant (NUHSRO/2014/063/SU/01), National University Health System Bridging Funds (NUHSRO/2017/023/Bridging/05), Ministry of Education Academic Research Fund Tier 1 (T1-NUHS Joint Grant Call FY17-2nd call-13), Lee Foundation Singapore (Single Cell Microfluidic Platform for Cardiovascular Disease) and Genome Institute of Singapore (SC18/22-1201 AR).

## Data availability

All genetic data are housed in the RAPTOR genomics repository (https://raptor.gisapps.org/). The data that support the findings of our study are available from the corresponding author of the study upon reasonable request.

## Supplementary Table legends

Supplementary Table 1: Clinical characteristics of patients with coronary artery disease included in the study

Supplementary Table 2: List of clonal hematopoiesis of indeterminate potential (CHIP)-associated genes included in the somatic mutation analysis

Supplementary Table 3: Distribution of genes harbouring somatic mutations in blood granulocytes and arterial tissues

Supplementary Table 4: Hallmark gene-set enrichment analysis of genes harbouring somatic mutations in arterial tissues

Supplementary Table 5: Sensitivity analysis of associations between somatic mutations and clinical phenotypes using a variant allele fraction (VAF) threshold of 2-25%

## Supplementary Figure legends

Supplementary Figure 1: Distribution of somatic mutations in CHIP-associated genes in blood granulocytes and arterial tissues at variant allele fraction (VAF) threshold of 2-25%.

## Notes

### Competing Interest Statement

The authors have declared no competing interest.

### Author Declarations

This study was approved by the National Healthcare Group (NHG) Domain Specific Review Board (NHG DSRB Reference Number: 2009/00216).

